# Normative Ranges for Wideband Middle Ear Muscle Reflex Magnitude: Limited Potential for Diagnosing Cochlear Deafferentation

**DOI:** 10.1101/2025.09.12.25335590

**Authors:** Naomi F. Bramhall, Garnett P. McMillan, Sean D. Kampel, Anne E. Heassler, Nicole K. Whittle, Haley A. Szabo

## Abstract

**Purpose:** Cochlear synaptopathy, the loss of the synapses between the inner hair cells and their auditory nerve fiber targets, is expected to be a common type of auditory deficit resulting from noise exposure or aging. Unfortunately, there is currently no means for diagnosing cochlear synaptopathy or other forms of cochlear deafferentation. Wideband middle ear muscle reflexes (MEMRs) have been proposed as a potential diagnostic indictor of cochlear deafferentation, but we lack normative ranges for MEMR magnitude. The objective of this study was to develop normative ranges for wideband MEMR magnitude that can be used to identify patients with abnormally weak MEMRs.

**Method:** Normative ranges were generated for ipsilateral and contralateral wideband MEMR magnitude in a population at low risk for cochlear synaptopathy due to young age, normal hearing thresholds, and minimal noise exposure history. The normative ranges were statistically adjusted for average distortion product otoacoustic emission (DPOAE) levels to account for possible impacts of outer hair cell dysfunction. To evaluate the ability of the normative ranges to differentiate between populations at low versus high risk of synaptopathy, measurements were also collected from military Veterans with normal hearing thresholds who reported at least one of the auditory complaints predicted to result from synaptopathy – tinnitus, speech perception in noise difficulty, or decreased sound tolerance.

**Results:** For individuals with poorer DPOAEs, it is not possible to fall below the lower bounds of the wideband MEMR normative ranges. For individuals with more robust DPOAEs, the lower bounds are very close to an MEMR magnitude indicating an absent reflex. Few individuals from the high-risk sample fell below the normative ranges, suggesting that these normative ranges do not identify significant cochlear deafferentation as expected.

**Conclusions:** Wideband MEMR magnitude normative ranges will not be effective as a stand-alone indicator of cochlear deafferentation.

## Introduction

Studies of human temporal bones suggest that age and noise-induced cochlear synaptopathy occur in humans (Wu et al., 2019; Wu et al., 2021), consistent with what is observed in animal models (e.g., Kujawa & Liberman, 2009; Sergeyenko et al., 2013). Cochlear synaptopathy is a type of cochlear deafferentation where the synapses between the inner hair cells and their afferent nerve fiber targets are lost, resulting in decreased peripheral input to the central auditory system. The predicted perceptual consequences of this type of auditory damage include tinnitus, hyperacusis, and difficulty with speech perception in noise (Kujawa & Liberman, 2015). However, because cochlear synaptopathy can only be confirmed through post-mortem temporal bone analysis, there is currently no means of diagnosing this condition in living humans.

Three auditory physiological measures that appear to be sensitive to cochlear synaptopathy have been identified in animal models. These include the amplitude of the first wave of the auditory brainstem response (ABR; Kujawa & Liberman, 2009; Sergeyenko et al., 2013), the envelope following response (EFR; Parthasarathy & Kujawa, 2018; Shaheen et al., 2015), and the middle ear muscle reflex (MEMR; Bharadwaj et al., 2022; Valero et al., 2018). ABR wave I amplitude is a measure of the synchronous firing of the auditory nerve fibers in response to a brief stimulus (see Bramhall, 2021 for a review of ABR wave I amplitude as an indicator of synaptopathy). The EFR measures the fidelity with which various elements of the auditory pathway (from the cochlea to the cortex) can code an amplitude modulated stimulus (see Van Der Biest et al., 2023 for a review of the EFR as an indicator of synaptopathy). The MEMR is a bilateral change in middle ear compliance evoked by high intensity sounds (see Trevino et al., 2023 for a review of the MEMR as a measure of synaptopathy). All three measures can be obtained non-invasively in living humans. Note that these physiological measures are general indicators of the degree of cochlear deafferentation, rather than specific measures of cochlear synaptopathy, because they will also be impacted by other sources of deafferentation such as loss of inner hair cells (IHCs) or spiral ganglion cells (SGCs). However, these physiological measures are expected to be primarily impacted by synaptopathy rather than other types of deafferentation because age-related loss of cochlear synapses outpaces loss of IHCs and SGCs (Wu et al., 2019), cochlear synapses are more vulnerable to noise-induced damage than IHCs (Wu et al., 2021), and noise-induced cochlear synapse loss occurs prior to loss of SGCs (Kujawa & Liberman, 2009). One complication of using these measures as indicators of cochlear deafferentation is that they have the potential to be impacted by outer hair cell (OHC) dysfunction in addition to deafferentation because they are all sound-evoked responses that require cochlear processing. Previous human studies of synaptopathy have typically addressed this issue by limiting study participation to individuals with clinically normal hearing and statistically adjusting for either average pure tone thresholds or average distortion product otoacoustic emissions (DPOAEs).

Of these three physiological measures, the MEMR has several advantages that would facilitate its use as a clinical measure of cochlear deafferentation. The MEMR reflex pathway includes the cochlea, auditory nerve, cochlear nucleus, superior olivary complex, facial nerve nucleus, and stapedius muscle (see review by Mukerji et al., 2010). The ipsilateral and contralateral MEMR pathways are both activated in response to a high intensity sound, with the contralateral pathway crossing over at the superior olivary complex. Reduction in auditory nerve activity secondary to cochlear synapse loss (or other types of deafferentation) should reduce the magnitude of the MEMR-induced change in middle ear compliance. Animal models suggest that both the ipsilateral and contralateral MEMR are weakened by synaptopathy (Bharadwaj et al., 2022; Valero et al., 2018). The MEMR can be obtained more quickly than the ABR or EFR, with a test time of approximately 5-10 minutes depending on the number of frequencies and levels tested. Another advantage of the MEMR over the ABR and EFR as an indicator of cochlear deafferentation is that it is already part of the standard audiometric test battery. In addition, measured MEMR strength may be relatively resistant to the impacts of OHC dysfunction (at least in individuals with normal hearing thresholds) because MEMR threshold elevation for a tonal elicitor is only observed when thresholds are poorer than approximately 50 dB HL (Gelfand et al., 1990) and a study in rabbits suggested that the MEMR is generated by the tails of auditory nerve tuning curves (Borg et al., 1990), which should not be impacted by small degrees of OHC dysfunction. Note that the MEMR probe used in animal studies of synaptopathy is a wideband probe (a click or chirp; Bharadwaj et al., 2022; Valero et al., 2018), while the probe used for clinical assessment is a 226 Hz probe tone. The wideband probe has the advantage of providing information about the change in eardrum compliance resulting from the MEMR across a wide range of frequencies, while the tonal probe only provides information at 226 Hz. Conclusions resulting from MEMR measurements obtained with a wideband probe versus a tonal probe may differ (see Figure 1 from Bramhall et al., 2025 for an example). Because the wideband probe provides additional information across frequency, it may be superior to a tonal probe for assessing cochlear deafferentation (Trevino et al., 2023).

**Figure 1.**
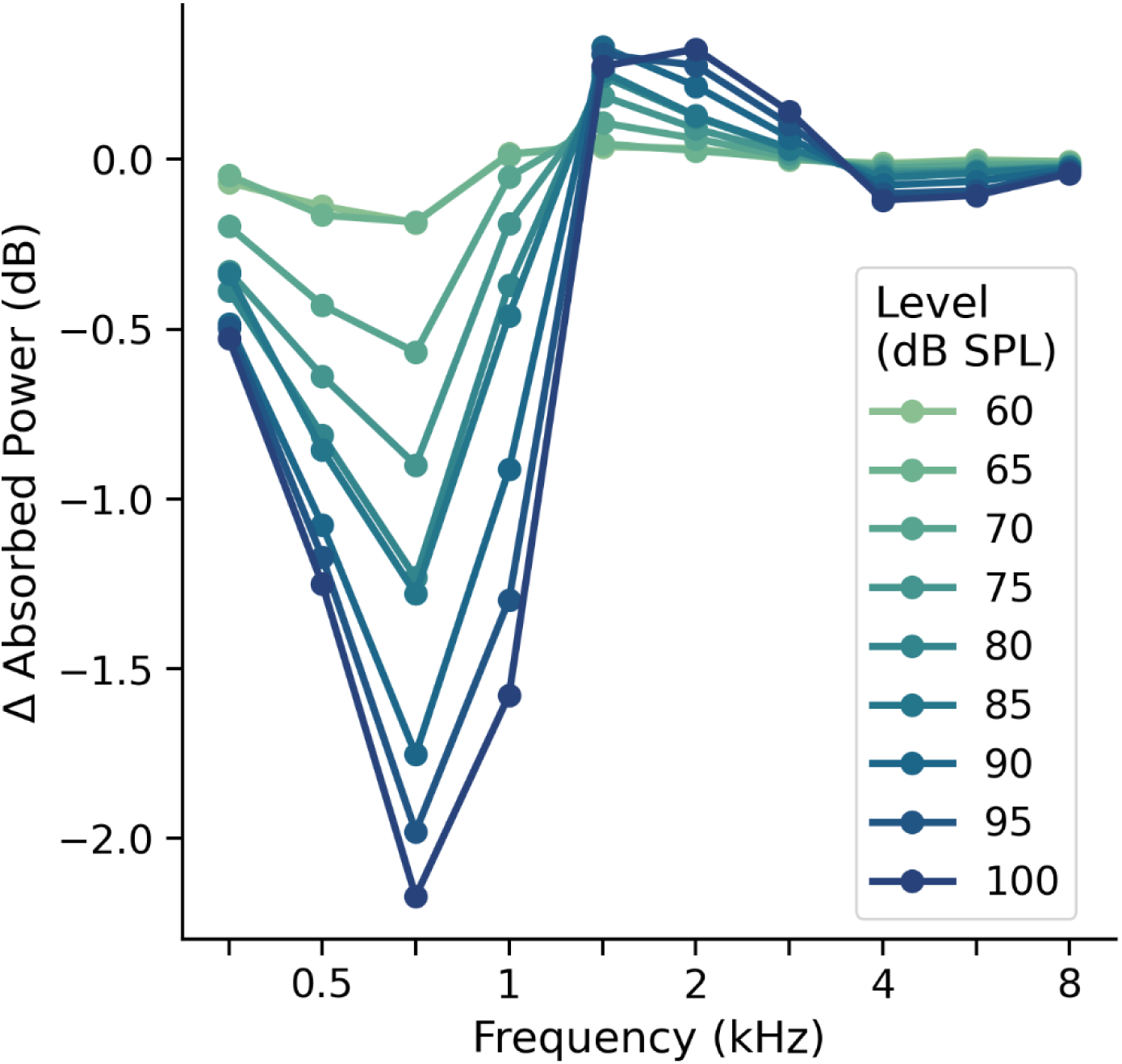
Example wideband MEMRs at increasing elicitor levels. Note that in this individual, the largest negative change in absorbed power between 0.25-2 kHz occurs at 0.75 kHz for each elicitor level. The MEMR magnitude (largest negative change in absorbed power from .25-2 kHz) for this individual would therefore be the change in absorbed power at 0.75 kHz for each elicitor level.

Human studies of cochlear deafferentation suggest that risk factors for synaptopathy (age and noise exposure) are associated with weaker wideband MEMRs, even when accounting for potential outer hair cell (OHC) dysfunction (Bharadwaj et al., 2022; Bramhall et al., 2022; Shehorn et al., 2020). In addition, in individuals with normal hearing thresholds, predicted perceptual consequences of synaptopathy (tinnitus and difficulty with speech perception in noise) are associated with weaker wideband MEMRs (Bramhall et al., 2023; Mepani et al., 2020; Wojtczak et al., 2017). This suggests that the wideband MEMR may be a useful diagnostic measure of cochlear deafferentation in humans. However, wideband MEMR cannot currently be used to identify deafferentation in individual patients because it is not clear what constitutes an abnormal MEMR measurement or how co-existing OHC dysfunction may impact the MEMR strength.

Because synaptopathy/deafferentation cannot be confirmed in living humans, we cannot objectively determine the MEMR strength that corresponds to a particular degree of deafferentation. An alternative is to measure MEMR strength in a large sample of individuals drawn from a population expected to be at low risk of synaptopathy/deafferentation and use their MEMR measurements to establish a normative range. Individuals who fall below these MEMR normative ranges would be expected to have significant cochlear deafferentation. Normative values are already commonly used for audiological assessment. For example, audiometric hearing thresholds are expressed in decibels hearing level (dB HL), which is referenced to normative values established in young adults assumed to have normal hearing (Fletcher & Wegel, 1922).

The objective of this study was to develop wideband ipsilateral and contralateral MEMR normative ranges in a population at low risk for cochlear synaptopathy due to their young age (18-35 years), minimal noise exposure history, normal hearing thresholds, and lack of auditory complaints (tinnitus, decreased sound tolerance, and difficulty with speech perception in noise). In addition, to account for the potential confounding impacts of OHC dysfunction, the normative ranges were adjusted for average DPOAE levels. The resulting normative ranges were then evaluated for their ability to identify individuals drawn from a population at high risk for synaptopathy (military Veterans with normal audiograms and auditory complaints) as having abnormally small MEMR magnitudes. The assumption was that if wideband MEMR magnitude is a good indicator of cochlear deafferentation, a higher percentage of the high-risk population would fall below the normative range than the low-risk normative population.

## Methods

### Participants

Participants were recruited from the Portland VA Health Care System, local colleges and universities, and the greater Portland, OR region. The Institutional Review Board of the VA Portland Health Care System gave ethical approval for this work. Informed consent was obtained from all participants prior to initiating any study activities, and participants were compensated for their time.

### Normative low-risk sample

Data was collected from 142 young adults (aged 18-35 years; 42 males, 101 females) who were assumed to be at low risk for synaptopathy due to their young age, minimal noise exposure history, and lack of auditory complaints (tinnitus, decreased sound tolerance, or difficulty with speech perception in noise). Individuals with a history of military service or a history of any firearm use were not included. All participants had normal audiometric thresholds (≤ 20 dB from 0.25-8 kHz) in at least one ear (referred to as the test ear) and normal tympanograms (±50 daPa, compliance 0.3-1.5 ml). Note that eleven low-risk participants had a normal tympanogram only in one ear. For these participants, only ipsilateral MEMR data was used in the analysis (with the stimulus/probe presented to the ear with the normal tympanogram). As a result, contralateral MEMR data from 131 low-risk participants was included in the analysis.

All participants from the normative low risk sample completed the Lifetime Exposure to Noise and Solvents Questionnaire (LENS-Q; Griest-Hines et al., 2021). The LENS-Q was scored as described in Griest-Hines et al. Previous work indicated a mean LENS-Q score of 4.1 for young non-Veterans with normal hearing and minimal noise exposure with a standard deviation of 0.8 (Bramhall et al., 2021). Accordingly, potential participants with LENS-Q scores ≥ 5 were excluded due to greater than minimal self-reported noise exposure.

Low-risk participants also completed a questionnaire that included questions about auditory complaints. Perception of tinnitus was assessed with the following question: “Have you experienced ringing, roaring, or buzzing in the ears or head (tinnitus) that lasts for at least 5 minutes?” Potential participants who reported tinnitus were excluded. Self-reported difficulty hearing speech in background noise was determined by asking participants to rate their perceived difficulty hearing in several situations (taken from the Hearing section of the Tinnitus and Hearing Survey [THS; Henry et al., 2015]). These situations include noisy or crowded places, understanding what people are saying on TV or in movies, understanding people with soft voices, and understanding what is being said in group conversations. Participants who reported difficulty in two or more of these situations were excluded. Participants were also asked the following question from the Sound Tolerance section of the THS: “Over the last week, sounds were too loud or uncomfortable for me when they seemed normal to others around me.” If they responded “yes” to this question, they were asked to provide two examples of sounds that were too loud or uncomfortable for them. If the examiner (a licensed audiologist) judged that their responses were consistent with decreased sound tolerance, they were excluded from the sample.

### High-risk comparison sample

For the purpose of comparing to the normative ranges, data was collected from a sample of 83 military Veterans, aged 23-49 years (52 males, 31 females), with normal audiograms and normal tympanograms in at least one ear who were expected to be at high risk of synaptopathy due to their likely history of military noise exposure and their report of at least one auditory complaint (tinnitus, decreased sound tolerance, or difficulty with speech perception in noise). Eleven Veteran participants had a normal tympanogram in only one ear. In these cases, only ipsilateral MEMR data from the ear with the normal tympanogram was used in the analysis. Accordingly, contralateral MEMR data was used from only 72 Veterans.

Veterans were asked the same questions about tinnitus, difficulty hearing speech in background noise, and decreased sound tolerance as the participants from the low-risk sample. Those that reported tinnitus, difficulty hearing speech in at least two of the situations from the THS and/or decreased sound tolerance (DST) were included in the high-risk sample. All Veterans also completed the Speech, Spatial and Qualities of Hearing Scale (SSQ12; Noble et al., 2013), a 12-item questionnaire that asks about difficulty hearing in a variety of situations. Average scores range from 0 to 10, with lower scores indicating greater difficulty. Veterans reporting tinnitus also completed the Tinnitus Functional Index (TFI; Meikle et al., 2012), a 25-item questionnaire that asks about the social and emotional effects of an individual’s hearing problem to assess the impact of hearing difficulty on their quality of life. Total scores range from 0 to 100, with higher scores indicating more severity.

## Procedures

### Distortion product otoacoustic emissions (DPOAEs)

DPOAE data were collected with a custom system consisting of an ER-10X probe microphone and EMAV software (Neely & Liu, 1993). A primary frequency sweep (DP-gram) from 1-16 kHz (1/3 octave intervals from 1-8 kHz and 1/6 octave intervals from 8-16 kHz) was obtained with a frequency ratio of *f_2_/f_1_*=1.2 and *L_1_/L_2_* levels of 65/55 dB forward pressure level (FPL). In-the-ear calibration was used to set *L_1_* and *L_2_* to the desired levels. An average of high frequency DPOAE levels (3-16 kHz or 3-8 kHz) was used to adjust the normative ranges for OHC function. Two different DPOAE averages were used because many DPOAE measurement systems are not capable of reliably measuring DPOAEs at frequencies above 8 kHz. DPOAE data was collected at each frequency according to the following parameters: until 48 seconds of artifact-free data were collected, until the noise floor was below -30 dB FPL, or the signal-to-noise ratio was ≥ 30 dB. If the measured DPOAE level at a particular frequency was less than the noise floor at that frequency, and the noise floor was low (< -20 dB FPL), the DPOAE level was set to -20 dB FPL. DPOAE levels that were below -20 dB FPL were not included in averages of DPOAE level across frequency.

The ER-10X enables FPL calibration, which increases the precision of the stimulus presentation level measurement, particularly for frequencies from 3-7 kHz in adult ear canals where interactions of incident and reflected waves produce pressure nodes or standing waves that can cause errors in standard sound pressure level (SPL) calibration (Konrad-Martin et al., 2016). Other than differences in the degree of measurement error, values in FPL and SPL are equivalent. Most commercially available DPOAE equipment provides the DPOAE measurements in dB SPL. Therefore, to avoid confusion when comparing the normative ranges to data collected from other pieces of equipment, DPOAE level is reported in dB SPL in all figures and tables in this report.

### Wideband middle ear muscle reflex (MEMR)

Ipsilateral and contralateral wideband MEMR measurements were obtained for each participant in a single ear (the test ear). Only the test ear (the ear where the elicitor stimulus was presented) was required to meet the audiometric threshold and tympanometric criteria for ipsilateral MEMR data to be included. However, for contralateral MEMR data to be included, the test ear had to meet the audiometric threshold criteria and both ears had to meet the tympanometric criteria. For contralateral measurements, the probe was placed in the non-test ear and the stimulus was presented in the test ear. If only one ear met the audiometric or tympanometric criteria, that ear was used as the test ear. If both ears met the criteria, but one ear had impacted cerumen or an ear piercing that might negatively impact testing, the other ear was tested. Otherwise, the test ear was selected by a rolling a die.

As described in Bramhall et al. (2025), wideband MEMR testing was conducted in a quiet room using an Interacoustics Titan probe coupled to a computer with a two-channel sound card and the Titan Research System software (Eden Prairie, MN). Five 95 dB peak-to-peak equivalent SPL clicks were presented to the probe ear with four noise burst elicitors presented to the stimulus ear in between each click. The probe and stimulus ear were the same for the ipsilateral condition. Broadband noise (BBN) low pass filtered at 8 kHz was used as the noise burst elicitor and was presented in 5 dB steps from 60 to 100 dB SPL, as measured by a Brϋel & Kjær (Darmstadt, Germany) head and torso simulator (Model 4128) connected to a Brϋel & Kjær LanXi digitizer.

However, artifact was noted in 2cc coupler recordings for the ipsilateral condition at elicitor levels above 85 dB SPL. Therefore, ipsilateral MEMR data for elicitor levels > 85 dB SPL were not included in the analysis. To increase scheduling flexibility, two different recording systems (Titan probe with Titan Research System software), located in two different rooms, were used to collect the MEMR data. One of these systems generates artifact consisting of pairs of 104 dB peak-to-peak equivalent SPL clicks (as measured with the Brϋel & Kjær head and torso simulator) in the stimulus/elicitor ear before presentation of the first contralateral elicitor stimulus for each intensity level and four clicks before the first ipsilateral elicitor for each level. The time between the last click and the first probe is approximately 4.5 s for the contralateral condition and 3 s for the ipsilateral condition. There is no click before the second elicitor stimulus. These clicks are not generated by the second recording system. We previously demonstrated that the median differences in contralateral MEMR magnitude between the two recording systems is very small and that there are no systematic differences between the systems (Bramhall et al., 2025).

As recommended by the manufacturer, the probe click was calibrated within 45 minutes of data collection by inserting the probe tip into four cylindrical acoustical propagation tubes measuring 7.94 mm in diameter (representative of the average adult ear). All MEMR measurements were collected at ambient pressure because all participants had clinically normal tympanograms and maintaining tympanometric peak pressure (TPP) was difficult in many participants. Schairer et al. (2022) showed that wideband MEMR thresholds (for a BBN elicitor) measured at ambient pressure in ears with normal tympanograms are similar or slightly lower than those measured at TPP.

Change in absorbed power relative to the first (baseline) click was calculated by the Titan Research System software as described in Keefe et al. (2017). Following Bramhall et al. (2025), the largest negative change in absorbed power occurring from 0.25-2 kHz was calculated so that MEMR strength could be expressed as a single value for each participant at each elicitor level (see **Figure 1**). This is similar to the approached used by Valero et al. (2018) in mouse. This value will be referred to as MEMR magnitude. Note that in cases where there was no MEMR in response to the elicitor, sometimes there was no negative change in absorbed power observed from 0.25-2 kHz. In these cases, the smallest change in absorbed power was used as the MEMR magnitude. These positive values were always very close to 0 dB (maximum of 0.04 dB).

### Generation of normative ranges

Normative ranges are often used to identify individuals with features that differ from those observed in a ‘healthy, normal’ population (Wright & Royston, 1999). A normative range represents specific percentiles of the measurement distribution for a normal reference population. For example, an 80% normative range covers the 10^th^ to 90^th^ percentiles of the measurement distribution for a normal population. Measurements that fall outside of the normative range are considered abnormal.

Conditional normative ranges vary based on specific patient characteristics, such as age, where measurements are collected from a large number of healthy individuals from different groups based on the characteristic (e.g., different age groups). Quantile regression is an alternative approach to generating conditional normative ranges that involves statistical modeling of the relationship between the population percentiles and a given characteristic. This approach fits regression models to estimate the 10^th^ and 90^th^ percentiles of the measurement distribution in a sample of healthy individuals with varying levels of the conditional variable (e.g., age). Unlike standard regression, which models the mean of the measurement distribution conditional on covariates, quantile regression models the percentiles of the measurement distribution conditional on covariates.

In this analysis, only data obtained from the low-risk sample contributed to the normative ranges. For each combination of MEMR condition (ipsilateral or contralateral) and elicitor level, the 10^th^ and 90^th^ percentiles of the MEMR magnitude distribution were conditioned on measured DPOAE levels averaged from either 3-16 kHz or 3-8 kHz.

Normative ranges were not calculated separately for males and females because previous studies have not shown any clear effect of sex on MEMR strength (Jerger et al., 1972; Osterhammel & Osterhammel, 1979).

## Results

### Ipsilateral MEMR normative ranges

Ipsilateral MEMR normative ranges adjusted for average DPOAE level from 3-16 kHz are shown in **Figure 2**. The normative ranges are plotted so that larger negative values of MEMR magnitude, indicating a stronger reflex, are at the upper bound of the normative range and smaller negative values of MEMR magnitude, indicating a small reflex or no reflex, are at the lower bound. At the lower elicitor levels (70 and 75 dB SPL), the lower bounds of the normative ranges essentially sit at an MEMR magnitude of 0 dB, indicating that even individuals with no ipsilateral MEMR at those elicitor levels would fall within the normative ranges. Note that a lower bound with a small positive value is also indicative of a lack of a reflex due to the absence of a negative peak. At the higher elicitor levels (80 and 85 dB SPL), the lower bounds of the normative ranges are 0 dB (or a small positive value) at lower average DPOAE levels (-15 to -10 dB SPL) and rise to progressively larger negative values for higher average DPOAE levels (up to -0.36 dB for an 85 dB SPL elicitor and an average DPOAE level of 10 dB SPL). **Figure 3** shows ipsilateral MEMR normative ranges adjusted for average DPOAE level from 3-8 kHz. These normative ranges are similar in shape to those shown in **Figure 2**, but with slightly smaller negative values at the lower bounds (i.e., the lower bounds are closer to 0 dB). The values for the lower bounds of the ipsilateral MEMR normative ranges adjusted for average DPOAE level from 3-16 kHz or 3-8 kHz are shown in **Table 1** and **Table 2**, respectively.

**Figure 2.**
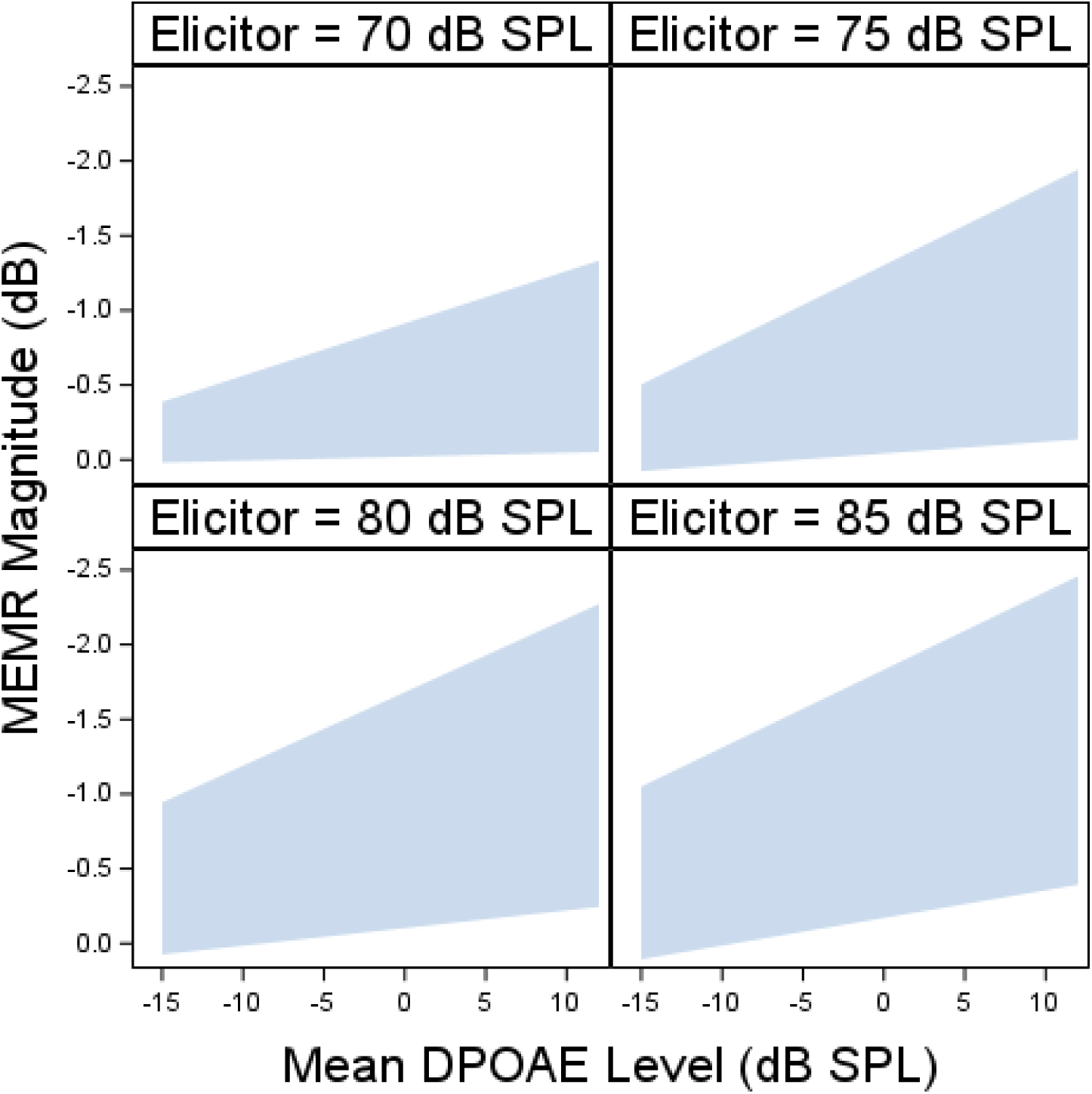
Ipsilateral wideband MEMR normative ranges adjusted for average DPOAEs from 3-16 kHz. Each panel shows the normative range for a different elicitor level. Blue shaded region indicates the DPOAE-adjusted normative range.

**Figure 3.**
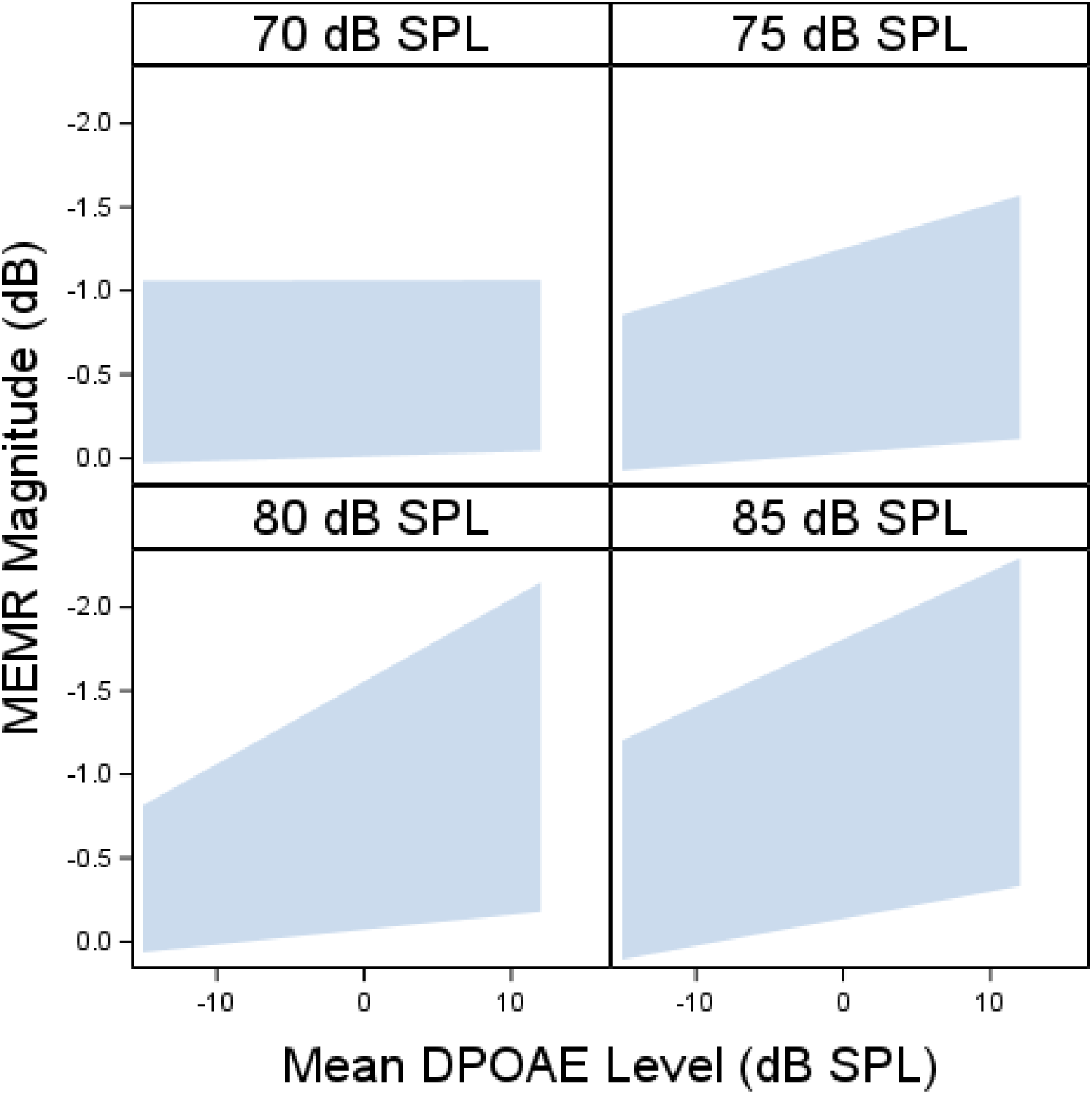
Ipsilateral wideband MEMR normative ranges adjusted for average DPOAEs from 3-8 kHz. Each panel shows the normative range for a different elicitor level. Blue shaded region indicates the DPOAE-adjusted normative range.

**Table 1.** Lower bounds for wideband MEMR normative ranges adjusted for average DPOAE levels from 3-16 kHz.

|  | Ipsilateral MEMR |  |  |  | Contralateral MEMR |  |  |  |  |
| --- | --- | --- | --- | --- | --- | --- | --- | --- | --- |
|  | Elicitor Level (dB SPL) |  |  |  |  |  |  |  |  |
| Average DPOAE level (dB SPL) | 70 | 75 | 80 | 85 | 80 | 85 | 90 | 95 | 100 |
| -15 | 0.02 | 0.08 | 0.07 | 0.11 | 0.05 | 0.02 | 0.19 | 0.08 | -0.01 |
| -10 | 0.01 | 0.04 | 0.02 | 0.01 | 0.01 | -0.02 | 0.09 | -0.04 | -0.12 |
| -5 | -0.01 | 0 | -0.04 | -0.08 | -0.02 | -0.06 | -0.01 | -0.15 | -0.24 |
| 0 | -0.02 | -0.04 | -0.10 | -0.17 | -0.06 | -0.10 | -0.12 | -0.27 | -0.35 |
| 5 | -0.03 | -0.08 | -0.16 | -0.26 | -0.09 | -0.14 | -0.22 | -0.38 | -0.47 |
| 10 | -0.05 | -0.12 | -0.22 | -0.36 | -0.13 | -0.18 | -0.32 | -0.50 | -0.58 |

**Table 2.** Lower bounds for wideband MEMR normative ranges adjusted for average DPOAE levels from 3-8 kHz.

|  | Ipsilateral MEMR |  |  |  | Contralateral MEMR |  |  |  |  |
| --- | --- | --- | --- | --- | --- | --- | --- | --- | --- |
|  | Elicitor Level (dB SPL) |  |  |  |  |  |  |  |  |
| Average DPOAE level (dB SPL) | 70 | 75 | 80 | 85 | 80 | 85 | 90 | 95 | 100 |
| -15 | 0.03 | 0.07 | 0.06 | 0.11 | 0.06 | 0.05 | 0.17 | 0.12 | 0.15 |
| -10 | 0.02 | 0.04 | 0.02 | 0.02 | 0.03 | 0.01 | 0.08 | 0.01 | 0.01 |
| -5 | 0 | 0 | -0.03 | -0.06 | 0 | -0.03 | -0.01 | -0.11 | -0.13 |
| 0 | -0.01 | -0.03 | -0.07 | -0.14 | -0.03 | -0.08 | -0.10 | -0.22 | -0.27 |
| 5 | -0.02 | -0.06 | -0.12 | -0.22 | -0.06 | -0.12 | -0.19 | -0.33 | -0.42 |
| 10 | -0.04 | -0.10 | -0.16 | -0.30 | -0.10 | -0.16 | -0.28 | -0.45 | -0.56 |

### Contralateral MEMR normative ranges

Contralateral MEMR normative ranges adjusted for average DPOAE level from 3-16 kHz are shown in **Figure 4**. As was observed for the ipsilateral MEMR normative ranges, at the lower elicitor levels (80-85 dB SPL) the lower bounds of the contralateral MEMR normative ranges are very close to 0 dB, except at the highest average DPOAE levels (-0.18 dB at 85 dB SPL for an average DPOAE level of 10 dB SPL). At the higher elicitor levels (95 and 100 dB SPL), the lower bounds of the normative ranges are 0 dB (or a small positive value) at the lowest average DPOAE level and rise to – 0.58 dB at the highest average DPOAE level (for the 100 dB SPL elicitor). **Figure 5** shows contralateral MEMR normative ranges adjusted for average DPOAE level from 3-8 kHz. The shapes of these normative ranges are similar to those shown in **Figure 4**, although the values at the lower bounds are slightly closer to 0 dB. The values for the lower bounds of the contralateral MEMR normative ranges adjusted for average DPOAE level from 3-16 kHz or 3-8 kHz are shown in **Table 1** and **Table 2**, respectively.

**Figure 4.**
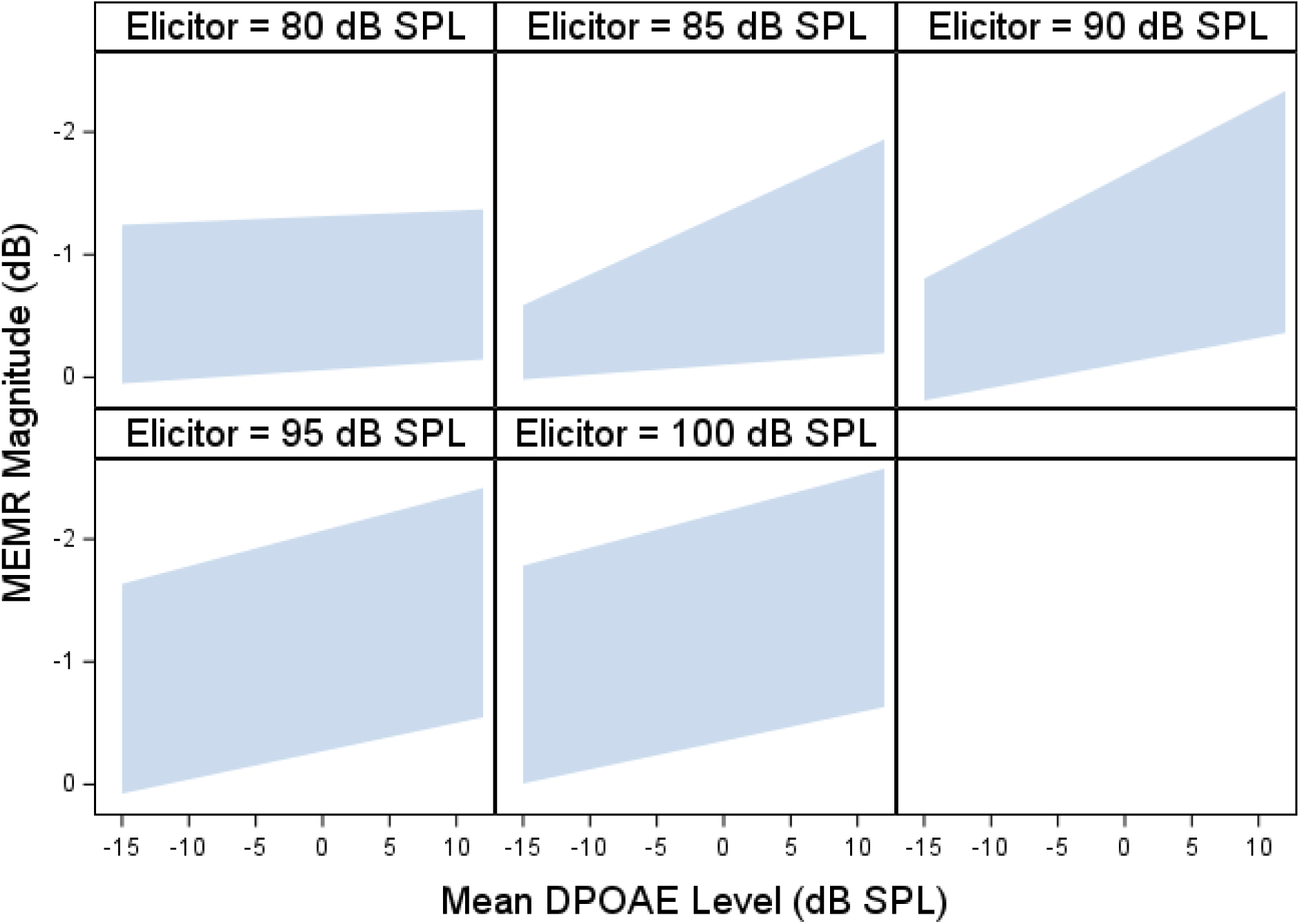
Contralateral wideband MEMR normative ranges adjusted for average DPOAEs from 3-16 kHz. Each panel shows the normative range for a different elicitor level. Blue shaded region indicates the DPOAE-adjusted normative range.

**Figure 5.**
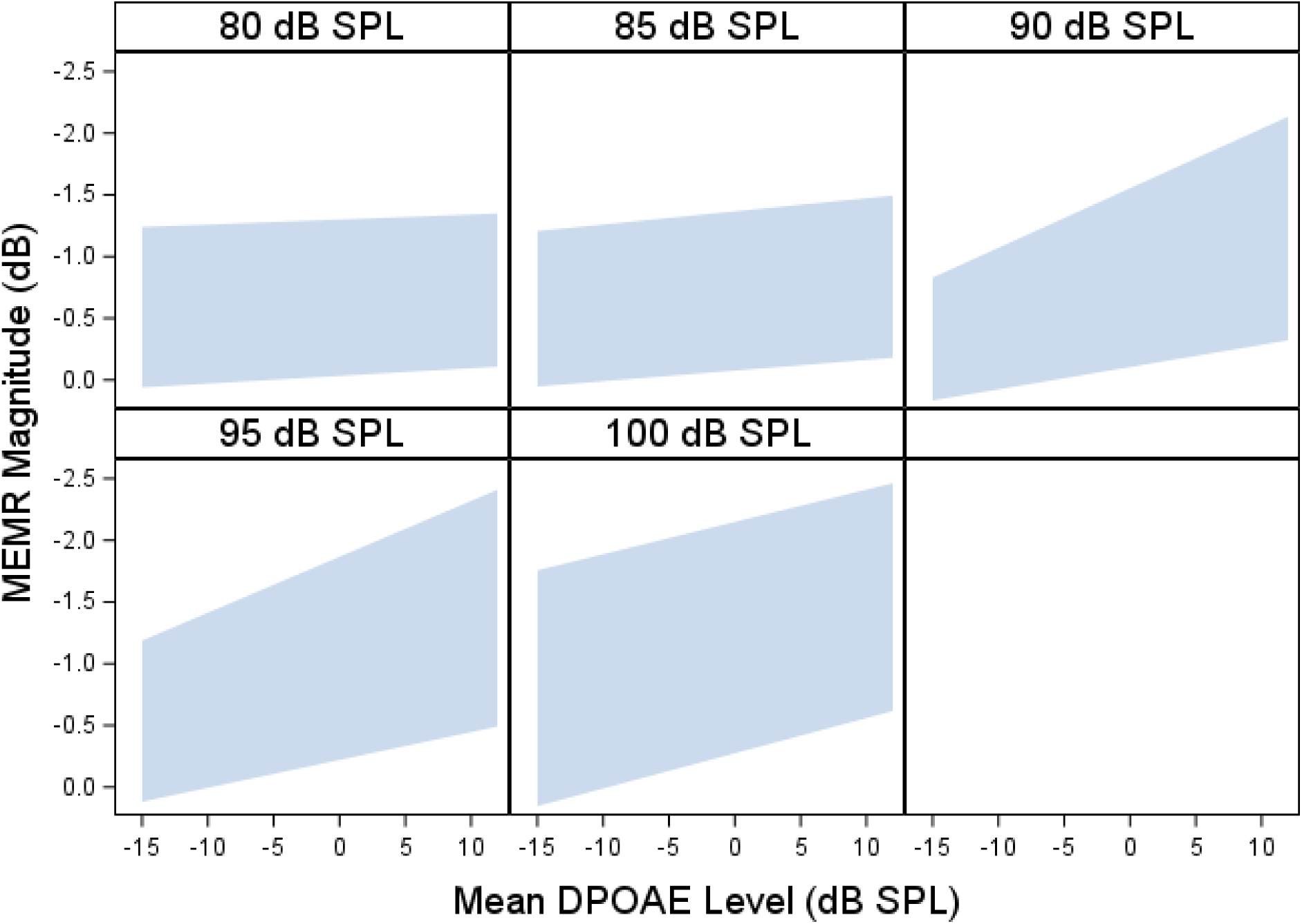
Contralateral wideband MEMR normative ranges adjusted for average DPOAEs from 3-8 kHz. Each panel shows the normative range for a different elicitor level. Blue shaded region indicates the DPOAE-adjusted normative range.

### Reporting of auditory complaints in the comparison sample at high risk for synaptopathy

**Table 3** summarizes the auditory complaints reported by the high-risk Veteran sample. The majority of Veterans reported more than one auditory complaint, with tinnitus and speech perception difficulty being the most common combination of complaints. On average, Veterans with more than one complaint reported greater tinnitus distress (as indicated by higher TFI scores) and speech perception difficulty (as indicated by SSQ12 score) than Veterans with only a single auditory complaint. Except for the complaint of decreased sound tolerance only, which was reported by a single male Veteran in his 20s, the mean age of Veterans reporting each complaint or combination of complaints ranged from 37-42 years.

**Table 3.** Report of auditory complaints in the high-risk sample.

| Complaint | # of males | # of females | Age in years | TFI score | SSQ score |
| --- | --- | --- | --- | --- | --- |
| Tinnitus only | 3 | 1 | 42 (5.8) | 20.1 (4.8) | 8.4 (0.6) |
| Speech perception only | 5 | 8 | 38.0 (5.6) | ----- | 6.7 (1.3) |
| DST only | 1 | 0 | In 20s | ----- | 7.6 |
| Tinnitus + speech perception | 31 | 12 | 37.9 (6.2) | 42.3 (15.6) | 6.0 (1.4) |
| Speech perception + DST | 3 | 1 | 39.5 (4.0) | ----- | 6.1 (1.5) |
| Tinnitus + speech perception + DST | 9 | 9 | 37.8 (6.9) | 45.0 (11.8) | 5.8 (1.5) |

The last 3 columns show mean values with the standard deviation in parentheses. DST – decreased sound tolerance; TFI – Tinnitus Functional Index; SSQ – Speech, Spatial, and Qualities of Hearing scale

### Comparison of high-risk sample to MEMR normative ranges

A small number of the Veterans from the high-risk sample fell below the MEMR normative ranges (**Figure 6** and **Figure 7**) when adjusted for DPOAE level from 3-16 kHz. At the highest elicitor levels (85 dB SPL for ipsilateral MEMR and 100 dB SPL for contralateral MEMR), 7.2% of the high-risk participants fell below the ipsilateral MEMR normative range and 6.3% fell below the contralateral MEMR normative range.

**Figure 6.**
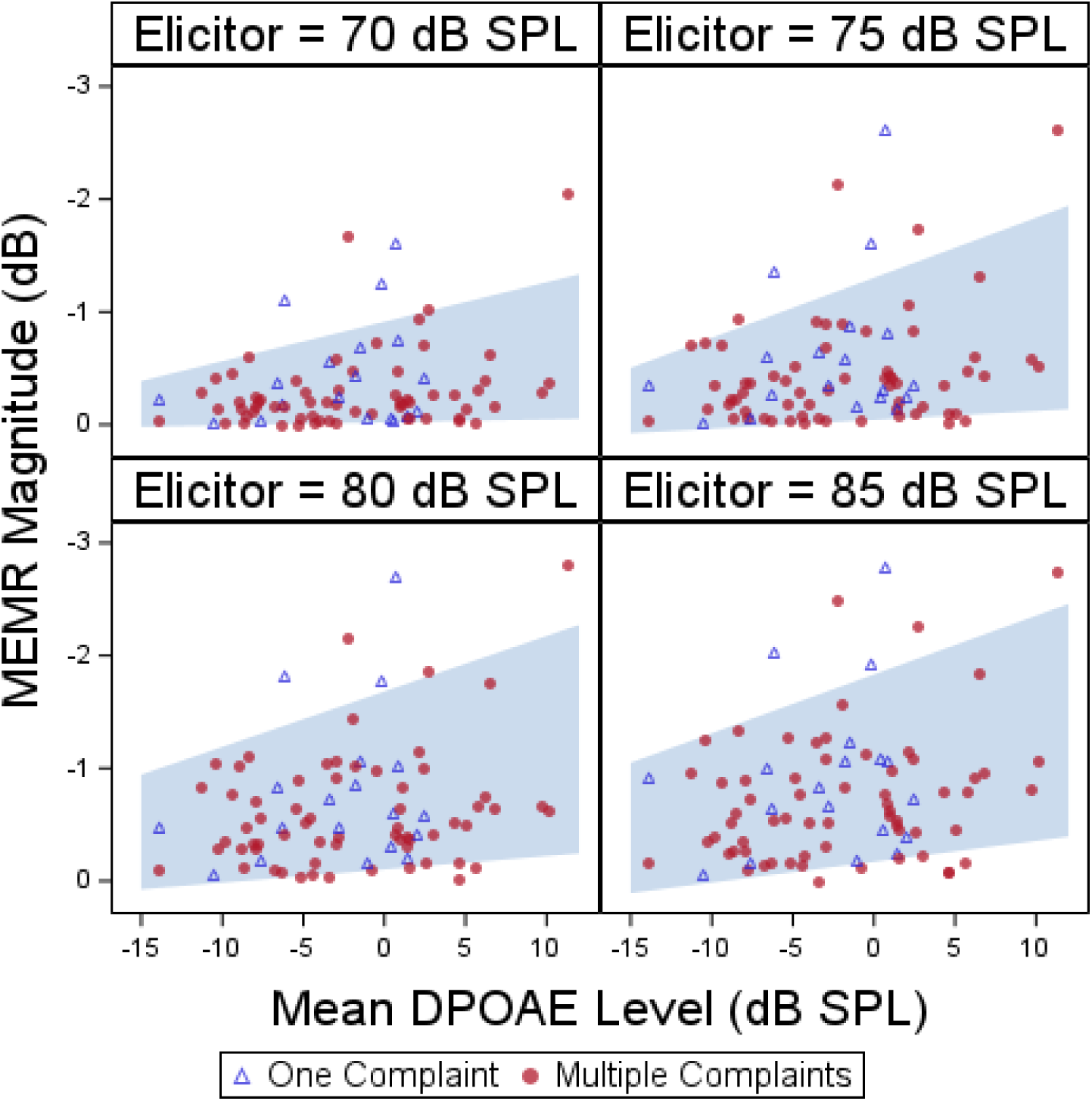
Comparison of high-risk sample to ipsilateral wideband MEMR normative ranges adjusted for average DPOAEs from 3-16 kHz. Each panel shows the normative range for a different elicitor level. Blue shaded region indicates the DPOAE-adjusted normative range. Symbols show data from individual high-risk Veterans with auditory complaints.

**Figure 7.**
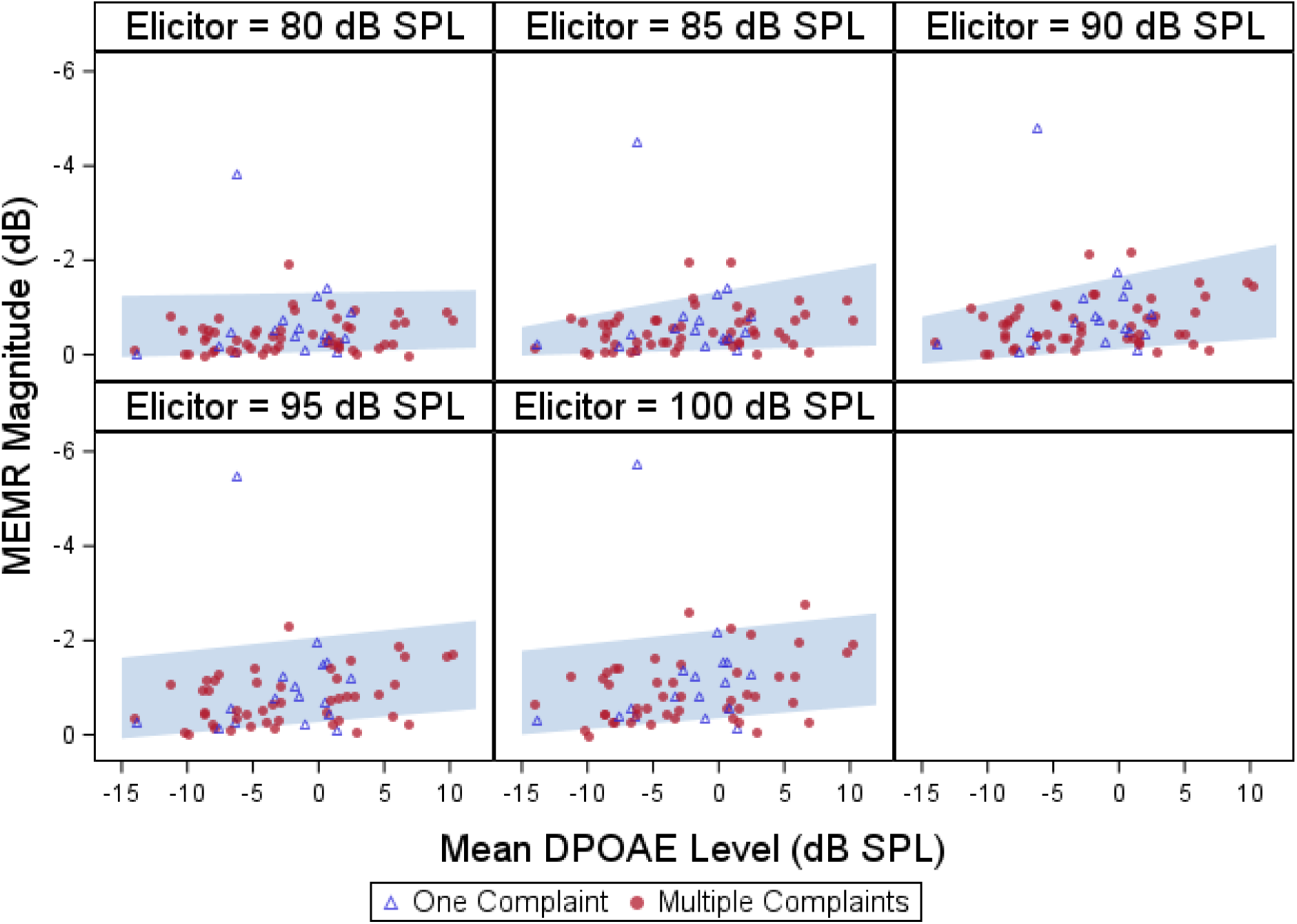
Comparison of high-risk sample to contralateral wideband MEMR normative ranges adjusted for average DPOAEs from 3-16 kHz. Each panel shows the normative range for a different elicitor level. Blue shaded region indicates the DPOAE-adjusted normative range. Symbols show data from individual high-risk Veterans with auditory complaints.

Veterans with more than one auditory complaint did not appear more likely to have lower MEMR magnitudes than Veterans with only a single auditory complaint. Several Veterans from the high-risk sample had MEMR magnitudes above the upper bound of the normative ranges. At the highest elicitor levels, 7.2% surpassed the ipsilateral MEMR normative range and 9.4% surpassed the contralateral MEMR normative range adjusted for average DPOAE level from 3-16 kHz. The percentage of high-risk Veterans falling above and below the ipsilateral and contralateral normative ranges adjusted for DPOAE level from 3-16 kHz are summarized in **Table 4**.

**Table 4.** Percentage of high-risk sample falling above or below MEMR normative ranges adjusted for average DPOAE level from 3-16 kHz.

|  | Ipsilateral MEMR |  |  |  | Contralateral MEMR |  |  |  |  |
| --- | --- | --- | --- | --- | --- | --- | --- | --- | --- |
|  | Elicitor Level (dB SPL) |  |  |  |  |  |  |  |  |
|  | 70 | 75 | 80 | 85 | 80 | 85 | 90 | 95 | 100 |
| Below normative range | 7.2% | 9.6% | 7.2% | 7.2% | 4.2% | 6.9% | 5.6% | 3.1% | 6.3% |
| Above normative range | 2.4% | 3.6% | 3.6% | 7.2% | 4.2% | 5.6% | 5.6% | 14.1% | 9.4% |

The percentage of high-risk participants falling outside the MEMR normative ranges at the highest elicitor levels when adjusting for DPOAE level from 3-8 kHz was very similar (7.2% and 7.8% below the ipsilateral and contralateral normative ranges, respectively and 7.2% and 9.4% above the ipsilateral and contralateral normative ranges). The percentage of high-risk Veterans falling above and below the ipsilateral and contralateral normative ranges adjusted for DPOAE level from 3-8 kHz are shown in **Table 5**.

**Table 5.** Percentage of high-risk sample falling above or below MEMR normative ranges adjusted for average DPOAE level from 3-8 kHz.

|  | Ipsilateral MEMR |  |  |  | Contralateral MEMR |  |  |  |  |
| --- | --- | --- | --- | --- | --- | --- | --- | --- | --- |
|  | Elicitor Level (dB SPL) |  |  |  |  |  |  |  |  |
|  | 70 | 75 | 80 | 85 | 80 | 85 | 90 | 95 | 100 |
| Below normative range | 6.0% | 7.2% | 6.0% | 7.2% | 4.2% | 5.6% | 5.6% | 4.7% | 7.8% |
| Above normative range | 1.2% | 4.8% | 3.6% | 7.2% | 4.2% | 5.6% | 4.2% | 12.5% | 9.4% |

## Discussion

### Comparison of ipsilateral versus contralateral MEMR normative ranges

There was no clear difference between the ipsilateral and contralateral MEMR normative ranges at the highest elicitor level in terms of the percentage of high-risk individuals who fell below the normative range. However, the highest elicitor level for the contralateral MEMR normative ranges (100 dB SPL) was considerably higher than the highest elicitor level for the ipsilateral MEMR normative ranges (85 dB SPL) due to problems with artifact in ipsilateral MEMR measurements obtained at 90-100 dB SPL. It’s possible that if reliable ipsilateral MEMR measurements had been collected at higher elicitor levels, normative ranges developed using those measurements would have resulted in more Veterans from the high-risk population falling below the normative range. Most previous studies that evaluated the use of wideband MEMR as an indicator of cochlear synaptopathy/deafferentation have only used either contralateral MEMR (Bramhall et al., 2022; Bramhall et al., 2023; Valero et al., 2018; Wojtczak et al., 2017) or ipsilateral MEMR (Bharadwaj et al., 2022; Casolani et al., 2022; Mepani et al., 2020). Shehorn et al. (2020) measured both ipsilateral and contralateral MEMR in the same sample but did not explicitly compare their relationships with risk factors and predicted perceptual consequences of deafferentation. For this reason, it is not clear if ipsilateral and contralateral wideband MEMR differ in terms of their sensitivity to deafferentation.

However, wideband MEMR magnitude at a particular elicitor level tends to be higher for ipsilateral than for contralateral elicitor presentation (Shehorn et al., 2020), increasing the likelihood of obtaining an ipsilateral reflex. For this reason, it is possible that the ipsilateral MEMR normative range for a 100 dB SPL elicitor would have a lower bound that was not as close to 0 dB as what is observed for the contralateral MEMR normative range for a 100 dB SPL elicitor, making it more effective at identifying individuals with deafferentation, particularly those with poorer DPOAEs.

### Impacts of DPOAE adjustment on MEMR normative ranges

Average DPOAE level had an impact on both the ipsilateral and contralateral MEMR normative ranges for higher elicitor levels, with poorer DPOAEs being associated with a lower bound near 0 dB and better DPOAEs being associated with a lower bound at slightly larger negative MEMR magnitudes. The upper bounds of the normative ranges also increased (i.e., values became larger negative numbers) as average DPOAE level increased. This suggests that poorer DPOAEs are associated with smaller MEMR magnitudes. It is not clear if this is because subclinical OHC dysfunction impacts MEMR magnitude or because individuals with poorer DPOAEs are just more likely to also have weaker MEMR magnitudes. However, the results of Bramhall et al. (2022) suggest that in non-Veterans with normal audiograms and minimal noise exposure history, poorer DPOAEs are not associated with smaller contralateral MEMR magnitudes. Factors such as middle ear dysfunction, noise exposure history, and age would be expected to impact both measurements. While all low-risk participants were young adults with minimal noise exposure history and had clinically normal tympanograms, variability in these factors would still be expected within the sample.

### Limited ability of MEMR normative ranges to identify significant cochlear deafferentation

Given that the MEMR normative ranges presented here are 80% normative ranges sampled from a population at low risk of synaptopathy, it would be expected that in a new sample from a low-risk population, roughly 10% of individuals would fall below the lower bound of the normative range and 10% would surpass the upper bound.

Because individuals from the high-risk population are expected to be more likely to have cochlear deafferentation than individuals from the low-risk population, if the MEMR was effective at detecting deafferentation, the high-risk individuals should be more likely than low risk individuals to fall below the normative range and less likely to surpass the normative range. However, that is not what is observed when comparing the high-risk sample to the normative ranges. For the ipsilateral MEMR normative ranges at the highest elicitor level (85 dB SPL) adjusted for DPOAE level from 3-16 kHz, 7.2% of the high-risk sample fell below the lower bound of the normative range (indicating an abnormally weak MEMR) and 7.2% surpassed the upper bound (indicating an unusually strong MEMR). Similarly, for the contralateral MEMR normative ranges at the highest elicitor level (100 dB SPL), 6.3% of the high-risk sample fell below the lower bound and 9.4% surpassed the upper bound. This suggests that either our assumption that individuals from the high-risk sample will have higher rates of cochlear deafferentation than the low-risk sample is not true, that the MEMR normative ranges are misclassifying as “normal” many individuals from the high-risk sample who have significant cochlear deafferentation, or that the MEMR measurements do not accurately reflect the reflex activity of the middle ear muscles. Considering that individuals with poorer DPOAEs cannot fall below the MEMR normative ranges because the lower bounds are at 0 dB, it seems reasonable to conclude that many individuals with significant cochlear deafferentation are not identified as abnormal by these normative ranges. While this is somewhat unexpected given previous studies that have shown mean differences in

MEMR magnitude between groups that differ in their risk for cochlear deafferentation due to age or noise exposure (Bharadwaj et al., 2022; Bramhall et al., 2022; Shehorn et al., 2020), the broad MEMR normative ranges suggest that wideband MEMR magnitudes are too variable in a low-risk population to effectively identify individuals likely to have significant cochlear deafferentation, particularly if they also have some OHC dysfunction. This interpretation is supported by the MEMR magnitude distributions for the low-risk and high-risk samples (**Figure 8**), which overlap considerably.

**Figure 8.**
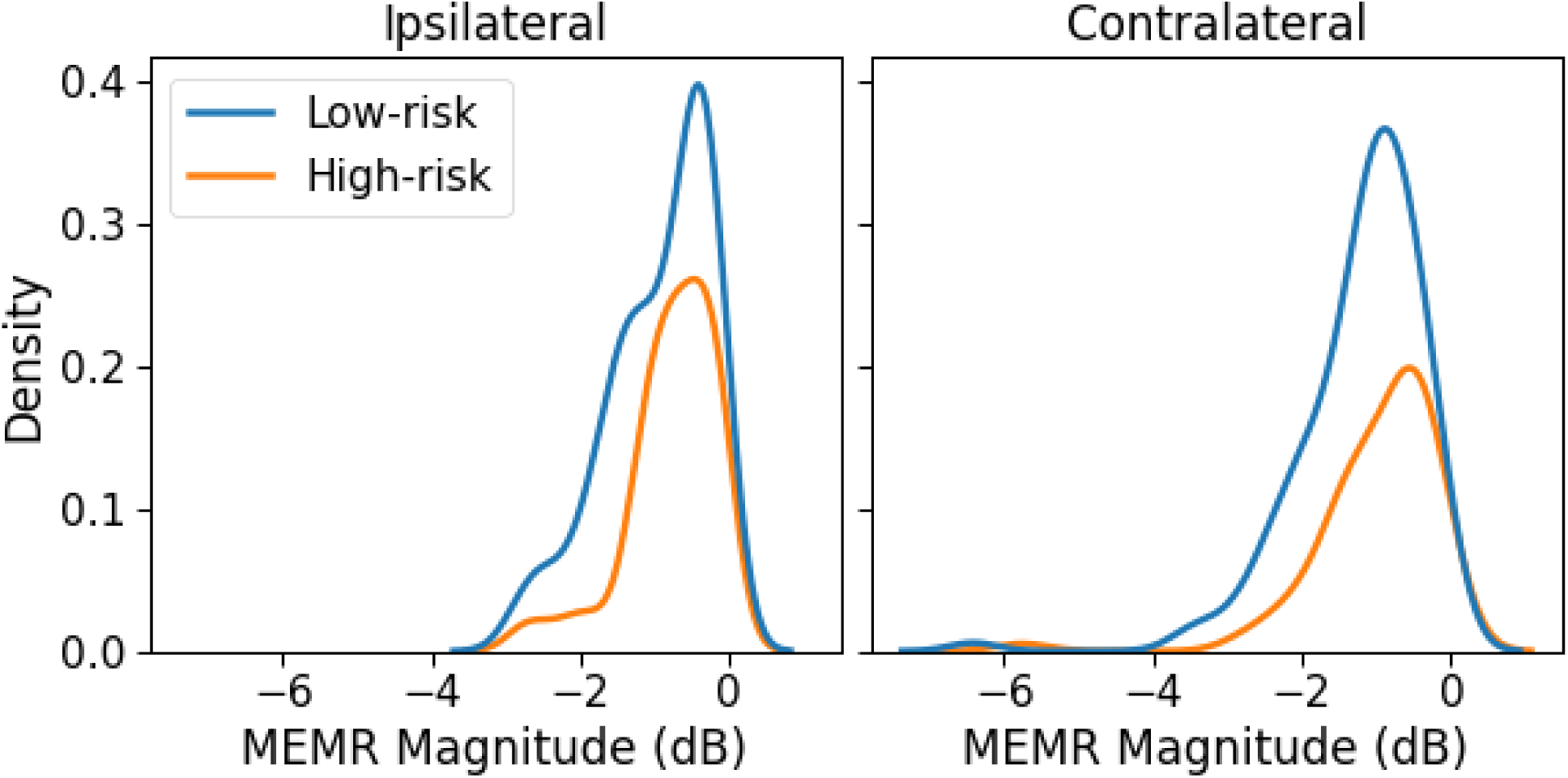
Wideband MEMR magnitude distributions for low-risk and high-risk samples. Kernel density estimation plots for ipsilateral and contralateral MEMR magnitude for the low-risk and high-risk samples.

### Future use of MEMR normative ranges

While it does not appear that wideband MEMR normative ranges alone will be effective at diagnosing significant cochlear deafferentation, these normative ranges could be used in the future as part of a diagnostic battery for cochlear deafferentation in individuals with robust DPOAEs. A high level (e.g., 100 dB SPL) normative range for wideband ipsilateral MEMR may be particularly useful for this purpose but would need to be developed using different equipment. Wideband MEMR testing can be completed in only a few minutes and falling below the wideband MEMR normative ranges in the context of normal audiometric thresholds and normal tympanometry suggests significant cochlear deafferentation. However, the diagnostic outcome for individuals falling within the normative ranges is inconclusive. These individuals would require additional testing to determine the likelihood of cochlear deafferentation.

## Conclusions

Ipsilateral and contralateral MEMR normative ranges were developed in a population at low risk for cochlear deafferentation due to their young age, normal hearing thresholds, and minimal noise exposure history. Unfortunately, due to the high variability in MEMR magnitudes in the normative sample, these normative ranges will not be informative as a stand-alone metric of deafferentation because it is only possible for individuals to fall below the normative ranges if they have robust DPOAEs. In addition, very few individuals from a Veteran population at high risk of cochlear deafferentation fell below the MEMR normative ranges, which would not be expected if the normative ranges were identifying individuals with significant cochlear deafferentation. Other physiological indicators of cochlear deafferentation, such as ABR wave I amplitude or EFR magnitude, may be better suited for diagnosing deafferentation.

## Data Availability

The datafiles generated during and/or analyzed during the current study are available from the corresponding author on reasonable request.

## Acknowledgements

This work was supported by the Department of Veterans Affairs, Veterans Health Administration, Rehabilitation Research and Development Service - Award #C3804-R/I01 RX003804 (to N.F.B.) and by resources and facilities at the VA National Center for Rehabilitative Auditory Research (NCRAR) [Center Award #C2361C/I50 RX002361] at the VA Portland Health Care System in Portland, OR. The opinions and assertions presented are private views of the authors and are not to be construed as official or as necessarily reflecting the views of the Department of Veterans Affairs.

## References

Bharadwaj, H. M., Hustedt-Mai, A. R., Ginsberg, H. M., Dougherty, K. M., Muthaiah, V. P. K., Hagedorn, A., . . . Heinz, M. G. (2022). Cross-species experiments reveal widespread cochlear neural damage in normal hearing. Communications Biology, 5(1), 733. doi:10.1038/s42003-022-03691-4

Borg, E., Counter, S. A., Engstrom, B., Linde, G., & Marklund, K. (1990). Stapedius reflex thresholds in relation to tails of auditory nerve fiber frequency tuning curves. Brain Research, 506(1), 79–84. doi:10.1016/0006-8993(90)91201-q

Bramhall, N. F. (2021). Use of the auditory brainstem response for assessment of cochlear synaptopathy in humans. Journal of the Acoustical Society of America, 150(6), 4440. doi:10.1121/10.0007484

Bramhall, N. F., McMillan, G. P., & Kampel, S. D. (2021). Envelope following response measurements in young veterans are consistent with noise-induced cochlear synaptopathy. Hearing Research, 408, 108310. doi:10.1016/j.heares.2021.108310

Bramhall, N. F., Reavis, K. M., Feeney, M. P., & Kampel, S. D. (2022). The Impacts of Noise Exposure on the Middle Ear Muscle Reflex in a Veteran Population. American Journal of Audiology, 1-17. doi:10.1044/2021_AJA-21-00133

Bramhall, N. F., Theodoroff, S. M., McMillan, G. P., Kampel, S. D., & Buran, B. N. (2023). Associations Between Physiological Correlates of Cochlear Synaptopathy and Tinnitus in a Veteran Population. Journal of Speech, Language, and Hearing Research, 66(11), 4635–4652. doi:10.1044/2023_JSLHR-23-00234

Bramhall, N. F., Whittle, N. K., Feeney, M. P., & McMillan, G. P. (2025). Test-Retest Differences in the Wideband Middle Ear Muscle Reflex. American Journal of Audiology, 34(1), 149–159. doi:10.1044/2024_AJA-24-00110

Casolani, C., Harte, J. M., & Epp, B. (2022). Categorization of tinnitus listeners with a focus on cochlear synaptopathy. PLoS One, 17(12), e0277023. doi:10.1371/journal.pone.0277023

Fletcher, H., & Wegel, R. L. (1922). The Frequency-sensitivity of Normal Ears. Proceedings of the National Academy of Sciences, 8(1), 5–6 2. doi:10.1073/pnas.8.1.5

Gelfand, S. A., Schwander, T., & Silman, S. (1990). Acoustic reflex thresholds in normal and cochlear-impaired ears: effects of no-response rates on 90th percentiles in a large sample. Journal of Speech and Hearing Disorders, 55(2), 198–205. doi:10.1044/jshd.5502.198

Griest-Hines, S. E., Bramhall, N. F., Reavis, K. M., Theodoroff, S. M., & Henry, J. A. (2021). Development and Initial Validation of the Lifetime Exposure to Noise and Solvents Questionnaire in U.S. Service Members and Veterans. American Journal of Audiology, 1–15. doi:10.1044/2021_AJA-20-00145

Henry, J. A., Griest, S., Zaugg, T. L., Thielman, E., Kaelin, C., Galvez, G., & Carlson, K. F. (2015). Tinnitus and hearing survey: a screening tool to differentiate bothersome tinnitus from hearing difficulties. American Journal of Audiology, 24(1), 66–77. doi:10.1044/2014_AJA-14-0042

Jerger, J., Jerger, S., & Mauldin, L. (1972). Studies in impedance audiometry. I. Normal and sensorineural ears. Archives of Otolaryngology-Head & Neck Surgery, 96(6), 513–523. doi:10.1001/archotol.1972.00770090791004

Keefe, D. H., Feeney, M. P., Hunter, L. L., & Fitzpatrick, D. F. (2017). Aural acoustic stapedius-muscle reflex threshold procedures to test human infants and adults. Journal of the Association for Research in Otolaryngology, 18(1), 65–88.

Konrad-Martin, D., Poling, G. L., Dreisbach, L. E., Reavis, K. M., McMillan, G. P., Lapsley Miller, J. A., & Marshall, L. (2016). Serial Monitoring of Otoacoustic Emissions in Clinical Trials. Otology & Neurotology, 37(8), e286–294. doi:10.1097/MAO.0000000000001134

Kujawa, S. G., & Liberman, M. C. (2009). Adding insult to injury: cochlear nerve degeneration after “temporary” noise-induced hearing loss. Journal of Neuroscience, 29(45), 14077–14085. doi:10.1523/JNEUROSCI.2845-09.2009

Kujawa, S. G., & Liberman, M. C. (2015). Synaptopathy in the noise-exposed and aging cochlea: Primary neural degeneration in acquired sensorineural hearing loss. Hearing Research, 330(Pt B), 191–199. doi:10.1016/j.heares.2015.02.009

Meikle, M. B., Henry, J. A., Griest, S. E., Stewart, B. J., Abrams, H. B., McArdle, R., . . . Vernon, J. A. (2012). The tinnitus functional index: development of a new clinical measure for chronic, intrusive tinnitus. Ear and Hearing, 33(2), 153–176. doi:10.1097/AUD.0b013e31822f67c0

Mepani, A. M., Kirk, S. A., Hancock, K. E., Bennett, K., de Gruttola, V., Liberman, M. C., & Maison, S. F. (2020). Middle Ear Muscle Reflex and Word Recognition in “Normal-Hearing” Adults: Evidence for Cochlear Synaptopathy? Ear and Hearing, 41(1), 25–38. doi:10.1097/AUD.0000000000000804

Mukerji, S., Windsor, A. M., & Lee, D. J. (2010). Auditory brainstem circuits that mediate the middle ear muscle reflex. Trends in Amplification, 14(3), 170–191. doi:10.1177/1084713810381771

Neely, S., & Liu, Z. (1993). *EMAV: Otoacoustic emission averager* (Tech Memo No. 17). Boys Town National Research Hospital.

Noble, W., Jensen, N. S., Naylor, G., Bhullar, N., & Akeroyd, M. A. (2013). A short form of the Speech, Spatial and Qualities of Hearing scale suitable for clinical use: the SSQ12. International Journal of Audiology, 52(6), 409–412. doi:10.3109/14992027.2013.781278

Osterhammel, D., & Osterhammel, P. (1979). Age and sex variations for the normal stapedial reflex thresholds and tympanometric compliance values. Scandinavian Audiology, 8(3), 153–158. doi:10.3109/01050397909076316

Parthasarathy, A., & Kujawa, S. G. (2018). Synaptopathy in the Aging Cochlea: Characterizing Early-Neural Deficits in Auditory Temporal Envelope Processing. Journal of Neuroscience, 38(32), 7108–7119. doi:10.1523/JNEUROSCI.3240-17.2018

Schairer, K. S., Putterman, D. B., Keefe, D. H., Fitzpatrick, D., Garinis, A., Kolberg, E., & Feeney, M. P. (2022). Automated Adaptive Wideband Acoustic Reflex Threshold Estimation in Normal-hearing Adults. Ear and Hearing, 43(2), 370–378. doi:10.1097/AUD.0000000000001102

Sergeyenko, Y., Lall, K., Liberman, M. C., & Kujawa, S. G. (2013). Age-related cochlear synaptopathy: an early-onset contributor to auditory functional decline. Journal of Neuroscience, 33(34), 13686–13694. doi:10.1523/JNEUROSCI.1783-13.2013

Shaheen, L. A., Valero, M. D., & Liberman, M. C. (2015). Towards a Diagnosis of Cochlear Neuropathy with Envelope Following Responses. Journal of the Association for Research in Otolaryngology, 16(6), 727–745. doi:10.1007/s10162-015-0539-3

Shehorn, J., Strelcyk, O., & Zahorik, P. (2020). Associations between speech recognition at high levels, the middle ear muscle reflex and noise exposure in individuals with normal audiograms. Hearing Research, 392, 107982. doi:10.1016/j.heares.2020.107982

Trevino, M., Zang, A., & Lobarinas, E. (2023). The middle ear muscle reflex: Current and future role in assessing noise-induced cochlear damage. Journal of the Acoustical Society of America, 153(1), 436. doi:10.1121/10.0016853

Valero, M. D., Hancock, K. E., Maison, S. F., & Liberman, M. C. (2018). Effects of cochlear synaptopathy on middle-ear muscle reflexes in unanesthetized mice. Hearing Research, 363, 109–118. doi:10.1016/j.heares.2018.03.012

Van Der Biest, H., Keshishzadeh, S., Keppler, H., Dhooge, I., & Verhulst, S. (2023). Envelope following responses for hearing diagnosis: Robustness and methodological considerations. Journal of the Acoustical Society of America, 153(1), 191. doi:10.1121/10.0016807

Wojtczak, M., Beim, J. A., & Oxenham, A. J. (2017). Weak Middle-Ear-Muscle Reflex in Humans with Noise-Induced Tinnitus and Normal Hearing May Reflect Cochlear Synaptopathy. eNeuro, 4(6), e0363–0317.2017 0361-0368. doi:10.1523/ENEURO.0363-17.2017

Wright, E. M., & Royston, P. (1999). Calculating reference intervals for laboratory measurements. Statistical Methods in Medical Research, 8(2), 93–112. doi:10.1177/096228029900800202

Wu, P. Z., Liberman, L. D., Bennett, K., de Gruttola, V., O’Malley, J. T., & Liberman, M. C. (2019). Primary Neural Degeneration in the Human Cochlea: Evidence for Hidden Hearing Loss in the Aging Ear. Neuroscience, 407, 8–20. doi:10.1016/j.neuroscience.2018.07.053

Wu, P. Z., O’Malley, J. T., de Gruttola, V., & Liberman, M. C. (2021). Primary Neural Degeneration in Noise-Exposed Human Cochleas: Correlations with Outer Hair Cell Loss and Word-Discrimination Scores. Journal of Neuroscience, 41(20), 4439–4447. doi:10.1523/JNEUROSCI.3238-20.2021

